# A cross-sectional questionnaire study: impaired awareness of hypoglycaemia remains prevalent in adults with type 1 diabetes and is associated with the risk of severe hypoglycaemia

**DOI:** 10.1101/2024.01.10.24301136

**Authors:** F. Baxter, N. Baillie, A. Dover, R.H. Stimson, F. Gibb, S. Forbes

## Abstract

**Objective:** Impaired awareness of hypoglycaemia (IAH) is a risk factor for severe hypoglycaemia (SH) in type 1 diabetes (T1D). Much of the IAH prevalence data comes from older studies where participants did not have the benefit of the latest insulins and technologies. This study surveyed the prevalence of IAH and SH in a tertiary adult clinic population and investigated the associated factors.

**Methods:** Adults (≥18 years) attending a tertiary T1D clinic completed a questionnaire, including a Gold and Clarke score. Background information was collected from health records.

**Results:** 189 people (56.1% female) with T1D (median [IQR] disease duration 19.3 [11.5, 29.1] years and age of 41.0 [29.0, 52.0] years) participated. 17.5% had IAH and 16.0% reported ≥1 episode of SH in the previous 12 months. Those with IAH were more likely to report SH (37.5% versus 11.7%, p=0.001) a greater number of SH episodes per person (median [IQR] 0 [0,2] versus 0 [0,0] P<0.001) and be female (72.7% versus 52.6%, p=0.036). Socio-economic deprivation was associated with IAH (p=0.032) and SH (p=0.005). Use of technology was the same between IAH vs aware groups, however, participants reporting SH were more likely to use multiple daily injections (p=0.026). Higher detectable C-peptide concentrations were associated with a reduced risk of SH (p=0.04).

**Conclusion:** IAH remains a risk factor for SH and is prevalent in females. Insulin pump and continuous glucose monitor use was comparable in IAH vs aware groups. Socioeconomic deprivation was associated with IAH and SH, making this an important population to target for interventions.

## Introduction

Type 1 diabetes (T1D) affects ∼35,000 people in Scotland (1). It is characterised by autoimmune destruction of the pancreatic beta cells leading, in time, to absolute or near absolute insulin deficiency (2). T1D is mainly managed with insulin replacement therapy which is given by multiple daily subcutaneous injections or continuous subcutaneous insulin infusion (CSII).

The landmark Diabetes Control and Complications (DCCT) trial found that the use of intensive insulin therapy in T1D reduced the risk of long-term microvascular complications, but that intensive therapy also increased the risk of hypoglycaemia (3). A recent prospective study identified hypoglycaemia as an ongoing burden for people with T1D who experience on average 73.3 hypoglycaemic events/patient-year (4). This effect is due in part to the inability of exogenous insulin to mimic the normal profile of endogenous insulin production, leading to relative insulin excess at inappropriate times (2), impairment of the normal compensatory hormone responses to lower blood glucose (5) and the loss of behavioural responses due to impaired awareness of hypoglycaemia (IAH) (2) which affects 20-40% of people with T1D (6, 7). IAH is a risk factor for severe hypoglycaemia (SH) (8), defined as an episode of hypoglycaemia requiring external assistance for recovery. IAH increases the risk of a SH event 6-fold (6, 9).

People with T1D can regain hypoglycaemia awareness through avoidance of hypoglycaemia (2, 10–12). Diabetes technologies such as continuous subcutaneous insulin infusion (CSII) and continuous glucose monitoring (CGM) can reduce overall episodes of hypoglycaemia (13, 14), improve glycaemic control and decrease the risk of microvascular complications (15). Advanced diabetes technologies, such as hybrid closed-loop systems, have been shown to reduce time in hypoglycaemia both in randomised controlled trials (RCTs)(16–19) and in real-world studies (20). However, their impact on IAH is not clear due to exclusion of participants with IAH from some RCTs (17) and other trials not reporting data on IAH (18). Additional studies investigating the effect of HCL systems on the counterregulatory response to hypoglycaemia and IAH in T1D are required (21) to further evaluate the possible benefits of these devices for those at risk of hypoglycaemia.

Technologies currently available in our clinic are: intermittently scanned CGM (isCGM), which users need to interact with in order to see their glucose data; real-time CGM (rtCGM) which transfers data in real-time to the user and CSII which can be used as part of a non-integrated system or as part of a hybrid closed-loop (HCL) system where there is automatic adjustment of insulin delivery based on readings from a rtCGM device. We surveyed an unselected population of adults with T1D to investigate if there has been a change in the prevalence of SH and IAH with the introduction of new technologies such as CGM and HCL systems. Health records of respondents were then screened to identify factors associated with IAH and SH.

## Methods

### Participants

Between the 1^st^ of July 2021 and the 31^st^ of August 2022 adults (≥18 years) attending a tertiary hospital T1D clinic in person were approached to complete the study survey. People with a diagnosis of T1D documented in their health record and a length of diagnosis of ≥2 years were considered eligible. Those unable to understand or complete the survey were excluded. The study was approved by the local research and development office (2021/0092) and research ethics committee (21/WA/0149). Written informed consent was obtained from participants.

### Questionnaire

The first part of the questionnaire included a Gold (9) and Clarke (22) Score. Both are validated methods for assessing hypoglycaemia awareness in people with T1D (23). In brief, for the Gold Score the participant is asked ‘Do you know when your hypos are commencing?’. They respond using a 7-point Likert scale with 1 indicating ‘always aware’ and 7 indicating ‘never aware’. A score of ≥4 represents IAH. The Clarke score comprises 8 questions that assess exposure to moderate and severe hypoglycaemia as well as assessing the glucose level for onset of symptoms. It gives a score of 0-8 with a score of ≥4 representing IAH.

The second part of the questionnaire collected additional information on employment status, education level, time off work or education due to hypoglycaemia, history of SH in the previous 12 months, driving status and use of diabetes technology.

Participant health records were reviewed to collect background information and demographic details including age, sex, age at diagnosis, HbA1c, insulin, date commenced CSII if applicable, date commenced intermittently scanned CGM (isCGM) or real time CGM (rtCGM) if applicable, postcode and hospital admissions in the previous 12 months related to diabetes. Socioeconomic status was assessed using the Scottish index for multiple deprivation (SIMD) quintile(24). C-peptide data was also collected from participant’s health records. Participant’s using an isCGM had a 2-week snapshot of their data collected from Libreview consisting of: time in range (TIR) 3.9-10 mmol/L, time below range (TBR) <3.9 mmol/L, time above range (TAR) >10 mmol/L, average glucose, standard deviation (SD) of glucose and coefficient of variation (CV) of glucose.

### C-peptide Analysis

C-peptide samples obtained prior to October 2021 were analysed by Abbot Architect and after this by Roche Elecsys. Values are reported down to the limit of detection, 3pmol/L for the Abbot system and 7pmol/L for the Roche system. Results below this limit of detection are reported as 0 pmol/L in this paper.

### Statistical Analysis

Results are reported as median (IQR) unless otherwise specified. Group differences in continuous variables were compared either using the unpaired t-test or Mann-Whitney U Test. Categorical variables were compared using the chi-square test. A p-value of <0.05 was considered significant. Statistical analysis was completed using IBM SPSS version 25.

## Results

*189 participants (56.1% female) completed the survey. IAH was defined as a Gold Score≥4, or where this was missing (2.1%), a Clarke score ≥4. The prevalence of IAH was 17.5%. Of note the prevalence of IAH was 17.5% using either score. When analysing respondents who completed both the Gold and Clarke questionnaires (93.1%), there was a significant positive correlation between the two scores, Pearson r 0.623 (P<0.001)*. 15.9% of respondents reported an episode of SH in the previous 12 months with a median (IQR) 0 (0,0) (range 0-12) number of episodes per person. The median (IQR) HbA1c was 60.0 (51.0, 67.0) mmol/mol (7.6 [6.8, 8.3]%). 56.6% of respondents were using multiple daily injections (MDI). 70.4% of respondents were using first generation insulin analogues as their bolus insulin and, of those using a basal insulin, 62.6% were using a second-generation analogue. The most common glucose monitoring method was isCGM with 81.0% of respondents using this. Of the 11.1% who were rtCGM users, 52.4% were using an unlicenced do-it-yourself (DIY) isCGM add-on to convert the device to a rtCGM sensor. Of the respondents using CSII, 9.8% were using hybrid closed loop (HCL) systems with 25% of these being a DIY HCL system.

Of the 144 isCGM users, Libreview data was available for 95 (66%). The median (IQR) TIR was 49 (34, 63)%, TBR 2 (0, 4)% and TAR 48 (33, 64)%. TIR was significantly positively correlated with the number of scans per day, Pearson *r* 0.4381 (P<0.0001) and significantly negatively correlated with HbA1c (*r* −0.7118, P<0.0001). There was also a significant negative correlation between TBR and HbA1c, *r* −0.3388 (p=0.0003). While TAR and HbA1c were positively correlated (*r* 0.7310, P<0.0001).

C-peptide data was available for 175 participants. The median (IQR) C-peptide was 3 (0, 16) pmol/L. C-peptide correlated significantly with the age at diagnosis, *r* 0.239 (p=0.001) and the diabetes duration, *r* ─0.398 (p<0.001). C-peptide did not significantly correlate with HbA1c (p=0.895), average glucose (p=0.254), TIR (p=0.473), TAR (p=0.363) and TBR (p=0.110).

42.6% of respondents came from the three most deprived SIMD quintiles. 5.9% were from the most deprived quintile 1, 19.1% from quintile 2 and 17.6% from quintile 3. The largest proportion of respondents, 36.7%, were from the least deprived SIMD quintile (quintile 5). Technology disparities existed between SIMD quintiles. MDI use was higher in the most deprived quintiles (1–3) compared to the least deprived (4–5), 66.3% versus 49.1% (p=0.025). Those from the three most deprived quintiles had a significantly higher median (IQR) HbA1c compared to those from the two least deprived quintiles, 64 (58, 75) mmol/mol (8 [7.4, 9)%] compared to 56 (50, 64) mmol/mol (7.3 [6.7, 8.0]%) (p<0.001).

72.7% of participants held a UK driving licence with 4.7% having previously surrendered their licence. While it did not reach statistical significance a numerically higher percentage of male respondents were drivers (77.1% versus 69.5%) and held a category of licence other than for driving a car alone (19.0% versus 12.7%).

Overall population characteristics are summarised in table 1.

**Table 1:**
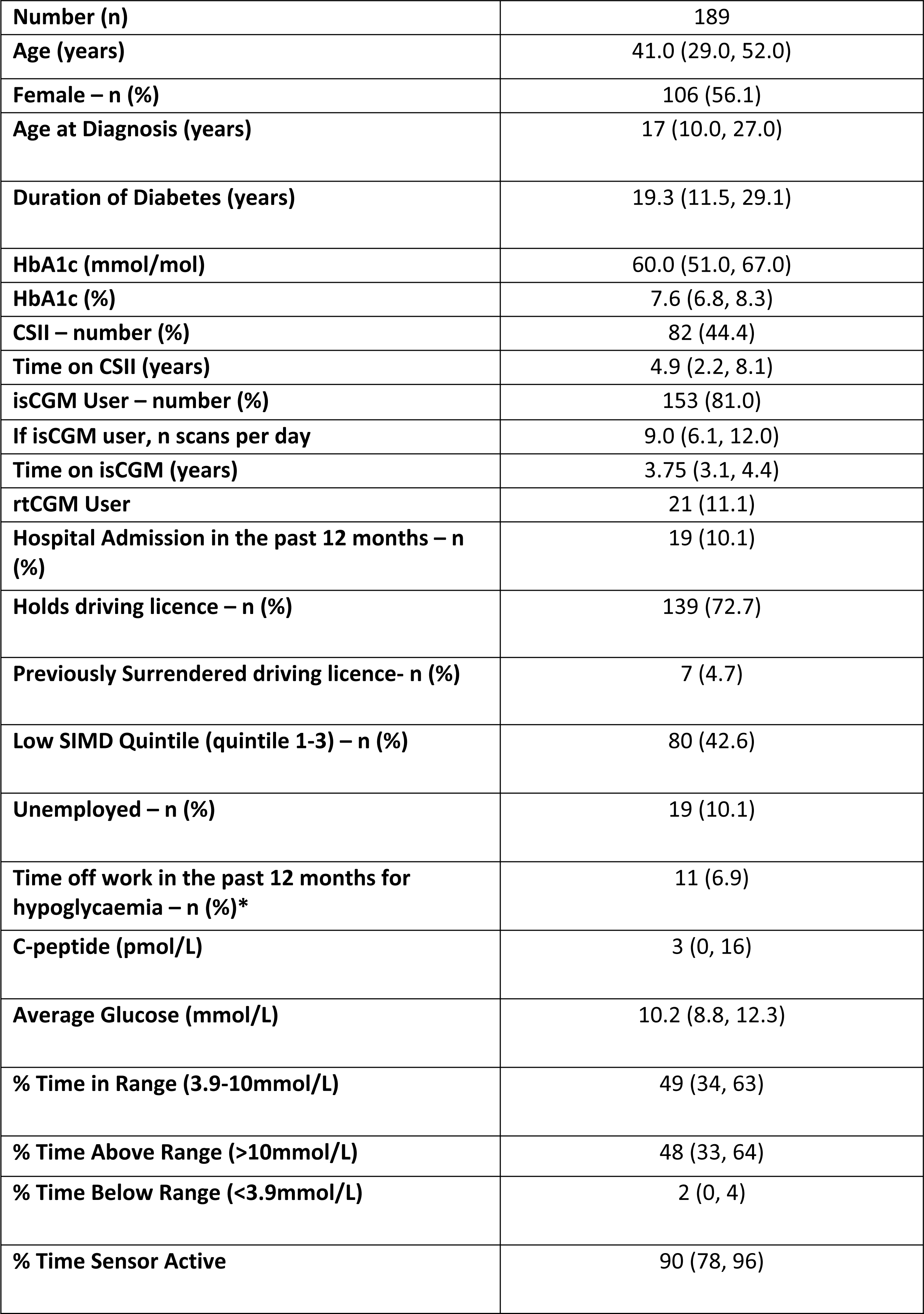
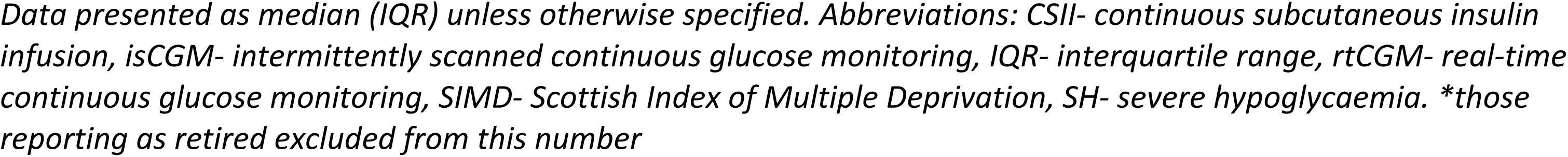
Overall Population Characteristics.

|  |  |
| --- | --- |
| **Number (n)** | 189 |
| **Age (years)** | 41.0 (29.0, 52.0) |
| **Female – n (%)** | 106 (56.1) |
| **Age at Diagnosis (years)** | 17 (10.0, 27.0) |
| **Duration of Diabetes (years)** | 19.3 (11.5, 29.1) |
| **HbA1c (mmol/mol)** | 60.0 (51.0, 67.0) |
| **HbA1c (%)** | 7.6 (6.8, 8.3) |
| **CSII – number (%)** | 82 (44.4) |
| **Time on CSII (years)** | 4.9 (2.2, 8.1) |
| **isCGM User – number (%)** | 153 (81.0) |
| **If isCGM user, n scans per day** | 9.0 (6.1, 12.0) |
| **Time on isCGM (years)** | 3.75 (3.1, 4.4) |
| **rtCGM User** | 21 (11.1) |
| **Hospital Admission in the past 12 months – n (%)** | 19 (10.1) |
| **Holds driving licence – n (%)** | 139 (72.7) |
| **Previously Surrendered driving licence- n (%)** | 7 (4.7) |
| **Low SIMD Quintile (quintile 1-3) – n (%)** | 80 (42.6) |
| **Unemployed – n (%)** | 19 (10.1) |
| **Time off work in the past 12 months for hypoglycaemia – n (%)<sup>\*</sup>** | 11 (6.9) |
| **C-peptide (pmol/L)** | 3 (0, 16) |
| **Average Glucose (mmol/L)** | 10.2 (8.8, 12.3) |
| **% Time in Range (3.9-10mmol/L)** | 49 (34, 63) |
| **% Time Above Range (>10mmol/L)** | 48 (33, 64) |
| **% Time Below Range (<3.9mmol/L)** | 2 (0, 4) |
| **% Time Sensor Active** | 90 (78, 96) |

Data presented as median (IQR) unless otherwise specified. Abbreviations: CSII- continuous subcutaneous insulin infusion, isCGM- intermittently scanned continuous glucose monitoring, IQR- interquartile range, rtCGM- real-time continuous glucose monitoring, SIMD- Scottish Index of Multiple Deprivation, SH- severe hypoglycaemia. \*those reporting as retired excluded from this number

### Impaired Awareness of Hypoglycaemia

IAH group differences are summarised in table 2. Participants with IAH compared to aware respondents were more likely to report an episode of SH in the previous 12 months, 37.5% compared to 11.7% (odds ratio [OR] 4.5 [95% confidence interval (CI) 2.0 to 10.9]) (p=0.001). The median (IQR) of SH episodes /person / year was higher in the IAH group, 0 (0, 2) compared to 0 (0, 0) (p<0.001) (figure 1A and 1B). Participants with IAH were also more likely to be female, 72.7% compared to 52.6% (p=0.036); older at the time of completing the survey, 44 (33, 61) years compared to 39.5 (29, 51) years (p=0.047); unemployed, 21.2% compared to 7.7% (p=0.005); and less likely to hold a driving licence, 56.3% compared to 76.1% (p=0.022). They were not more likely to have surrendered their driving licence in the past.

**Table 2:** Characteristics of Subgroups of Impaired Awareness of Hypoglycaemia (IAH)

| Subgroup | Normal Awareness | Impaired Awareness | P-Value |
| --- | --- | --- | --- |
| Number (n) – (%) | 156 (82.5) | 33 (17.5) |  |
| Age (years) – median (IQR) | 39.5 (29, 51) | 44 (33, 61.0) | **0.047** |
| Female – n (%) | 82 (52.6) | 24 (72.7) | **0.036** |
| Age at Diagnosis (years) – median (IQR) | 16.0 (10.2, 25.0) | 20.0 (10.0, 37.5) | 0.092 |
| Duration if Diabetes (years) – median (IQR) | 19.3 (11.9, 28.3) | 20.8 (9.6, 38.1) | 0.779 |
| HbA1c (mmol/mol) | 59.5 (51.0, 67.0) | 63.5 (50.3, 73.0) | 0.341 |
| HbA1c (%) | 7.6 (6.8, 8.3) | 8.0 (6.8, 8.8) |  |
| CSII User – n (%) | 69 (44.2) | 13 (39.4) | 0.701 |
| Time on CSII (years) | 5.3 (2.5, 8.2) | 2.3 (0.3, 6.8) | 0.121 |
| isCGM – n (%) | 128 (82.1) | 25 (75.8) | 0.591 |
| If isCGM user, n scans per day | 9 (6.5, 12.0) | 9 (6.0, 11.0) | 0.584 |
| Time on isCGM (years) | 3.7 (3.1, 4.4) | 3.9 (3.2, 4.4) | 0.905 |
| rtCGM User | 17 (10.9) | 4 (12.1) | 0.591 |
| SH in the past 12 months – n (%) | 18 (11.7) | 12 (37.5) | **0.001** |
| No. SH Episodes | 0 (0, 0) | 0 (0, 2) | **<0.001** |
| Hospital Admission in the past 12 months – n (%) | 13 (8.3) | 6 (18.2) | 0.087 |
| Holds driving licence – n (%) | 118 (76.1) | 18 (56.3) | **0.022** |
| Low SIMD Quintile (quintile 1-3) – n (%) | 60 (39.4) | 20 (60.6) | **0.032** |
| Unemployed – n (%) | 12 (7.7) | 7 (21.2) | **0.005** |
| Time off work in the past 12 months for hypoglycaemia – n (%)* | 8 (5.9) | 3 (12.5) | 0.374 |
| C-peptide (pmol/L) | 3 (0, 18.5) | 4.5 (0, 14.5) | 0.962 |
| Average glucose (mmol/L) | 10 (8.8, 12.1) | 11.9 (10.6, 14.7) | **0.041** |
| Time in range (3.9-10 mmol/L) | 50 (36, 64.5) | 35 (26.3, 46.3) | **0.031** |
| Time below range (<3.9mmol/L) | 2 (0, 4) | 1 (0, 3) | 0.161 |
| Time above range (>10 mmol/L) | 46 (32.5, 62) | 63.5 (52.5, 71) | **0.025** |
| % Time Sensor Active | 91 (79, 96.5) | 77 (62.5, 93.5) | 0.081 |

Data presented as mean (SD) unless otherwise specified. Abbreviations: CSII- continuous subcutaneous insulin infusion, isCGM- intermittently scanned continuous glucose monitoring, IQR- interquartile range, rtCGM- real-time continuous glucose monitoring, SIMD- Scottish Index of Multiple Deprivation, SH- severe hypoglycaemia

**Fig 1A.**
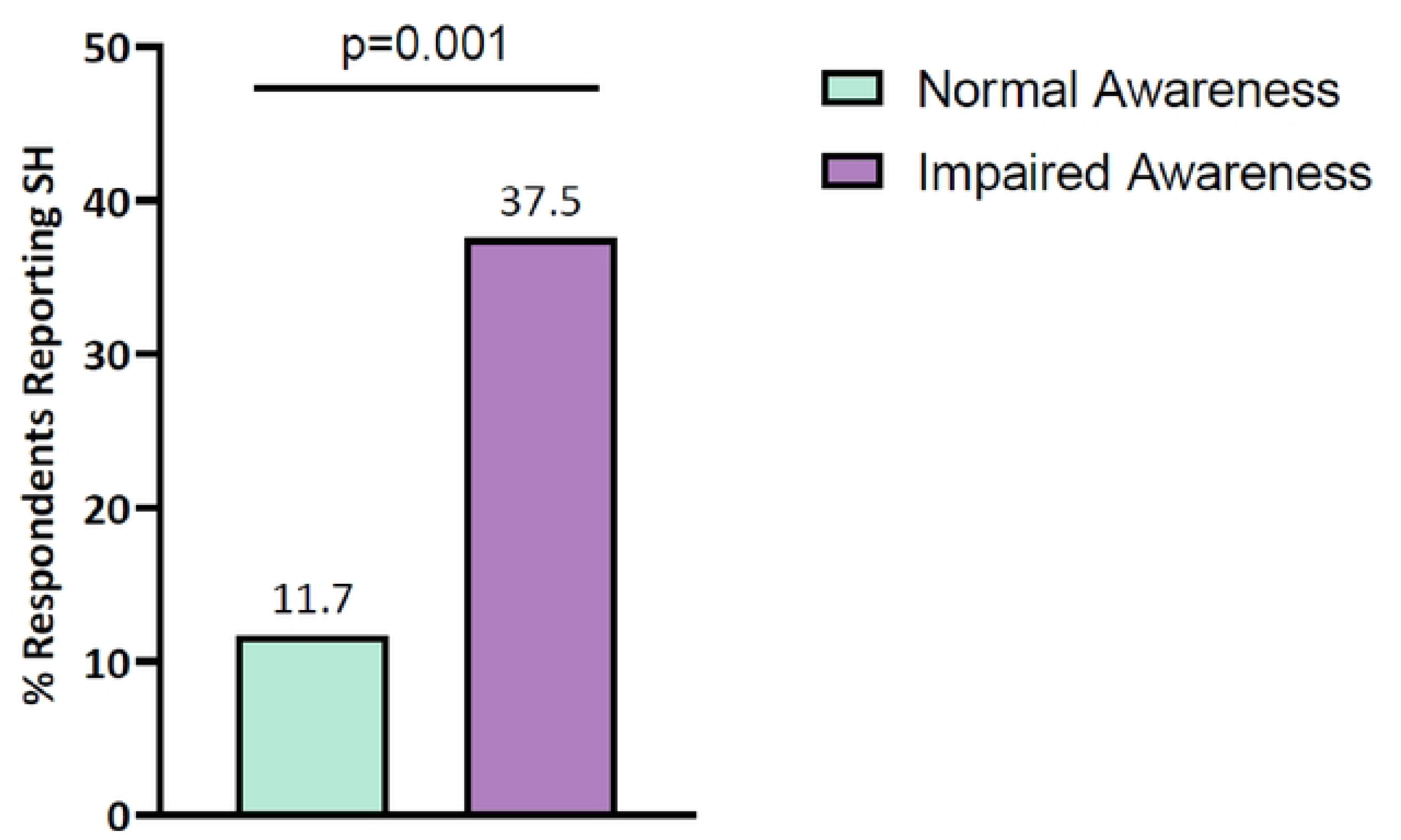
Respondents Reporting Severe Hypoglycaemia (SH) in the Previous 12 months.

**Fig 1B.**
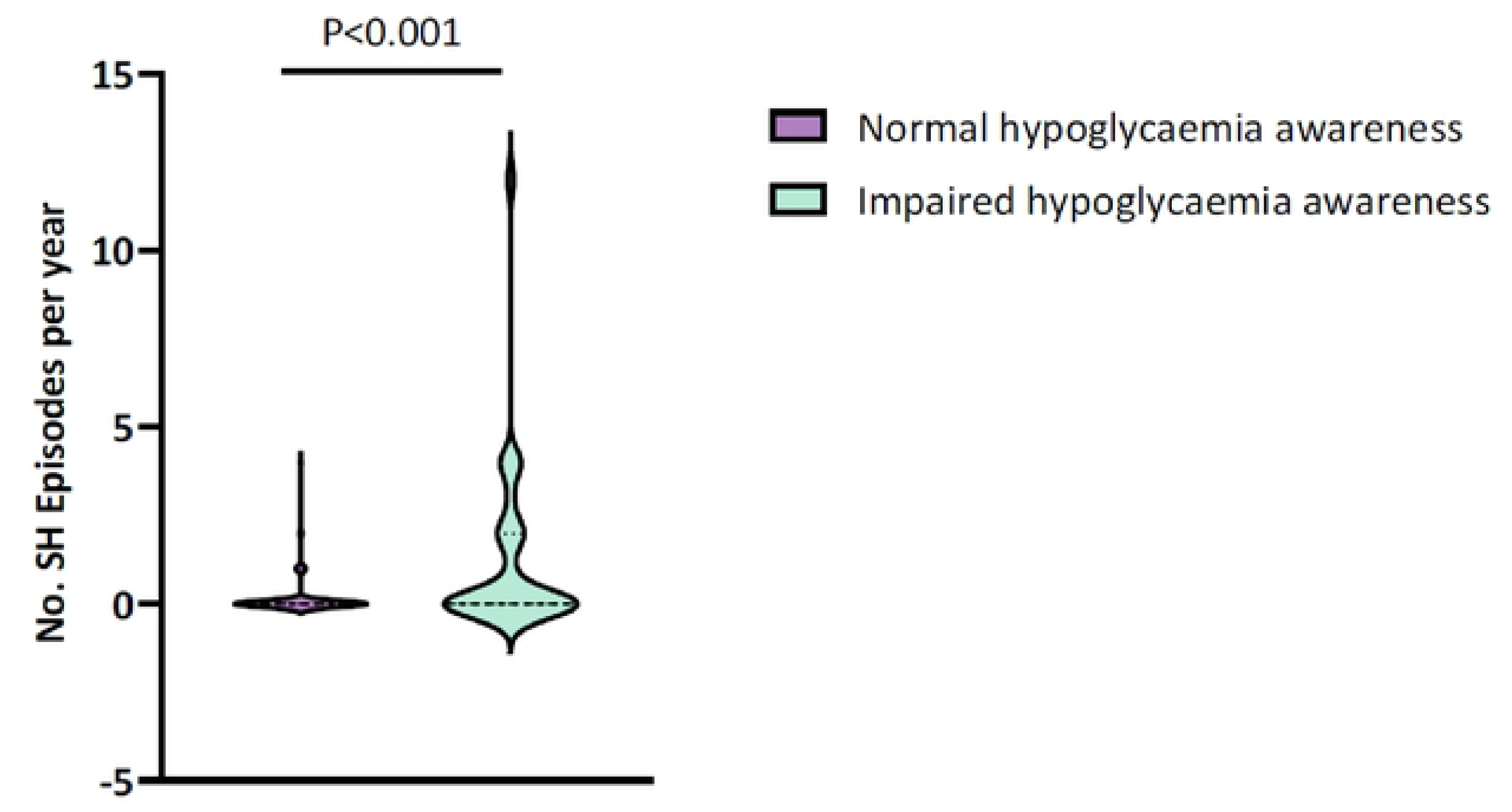
Number of SH events per patient in the previous year.

IAH was associated with socioeconomic deprivation, 60.6% of respondents with IAH were in SIMD quintile 1 to 3 compared to 39.4% of respondents with normal awareness (p=0.032) (figure 2). Numerically a higher percentage of people with IAH had a diabetes-related hospital admission in the previous 12 months, 18.2% vs. 8.3%, but this did not reach statistical significance (p=0.087). There was also a trend for people with IAH being diagnosed at an older age with a median (IQR) age at diagnosis of 20 (10.0, 37.5) years versus 16.0 (10.2, 25.0) years (p=0.092).

**Fig 2:**
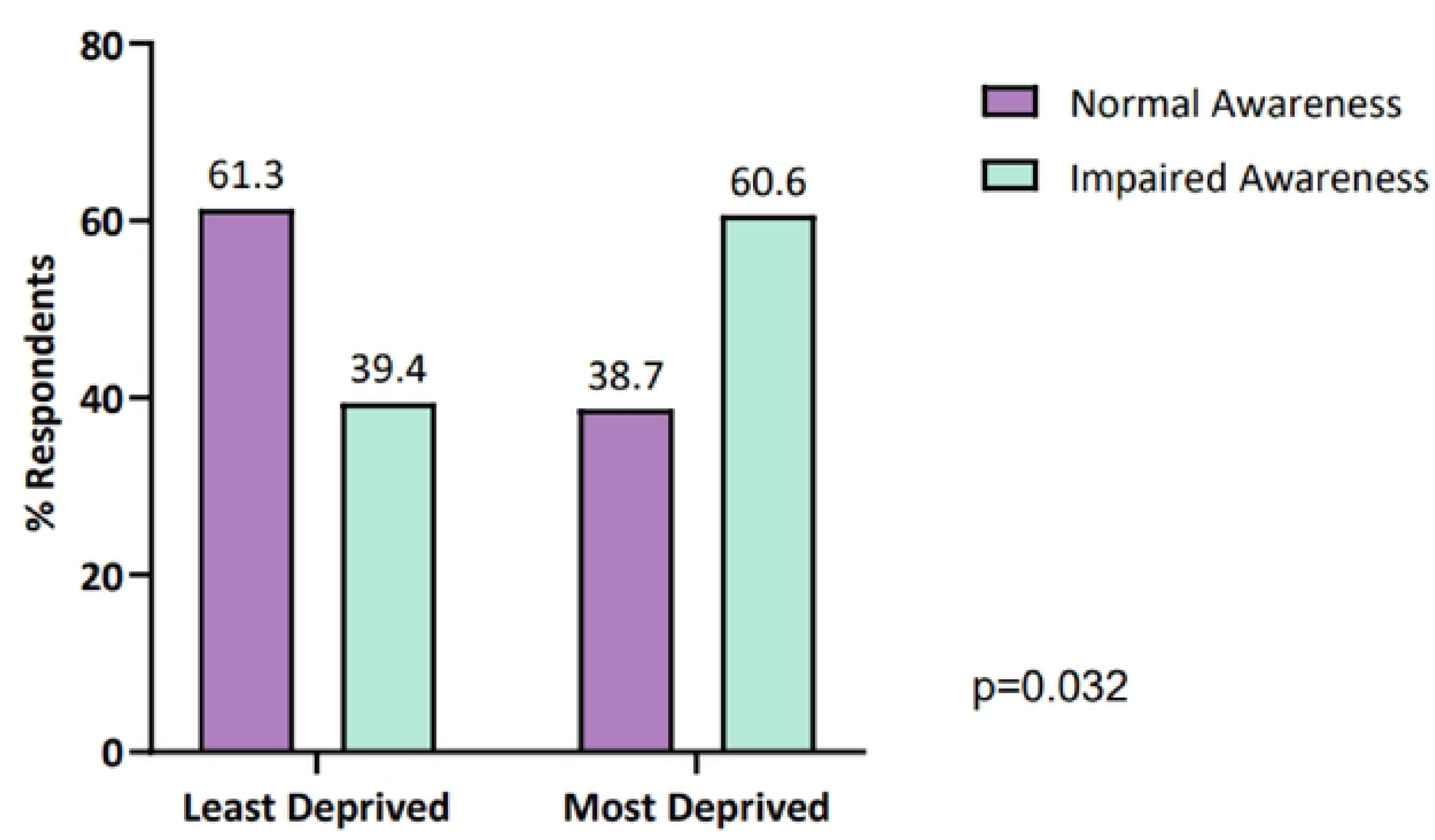
Impaired Awareness of Hypoglycaemia by SIMD Quintile Least deprived-quintile 4 and 5. Most deprived-quintile 1-3. Abbreviations: SIMD-Scottish index of multiple deprivation.

There was no group difference between respondents with impaired and normal awareness in the use of CSII (39.4% compared to 44.2% [p=0.701]), isCGM (75.8% compared to 82.1% [p=0.591]) or rtCGM. (12.1% compared to 10.9% [p=0.591]). More people with normal awareness were using a DIY isCGM add-on than in the IAH group, 64.7% compared to 0% (p=0.035).

There was no significant group difference in C-peptide levels between those with IAH vs aware, 4.5 (0, 14.5) pmol/L compared to 3 (0, 18.5) pmol/L (p=0.962)

Respondents reporting IAH had a higher average glucose, 11.9 (10.6, 14.7) mmol/L compared to 10 (8.8, 12.1) mmol/L (p=0.041); a lower TIR, 35% (26.3, 46.3%) compared to 50% (36, 64.5%) (p=0.031) and a higher TAR, 63.5% (52.5, 71%) compared to 46% (32.5, 62%) (p=0.025).

### Severe Hypoglycaemia

SH group differences are summarised in table 3. Participants reporting SH in the previous 12 months were compared to those with no history of SH and found to have: a higher median (IQR) HbA1c, 64.5 (55.7, 75.3) mmol/mol (8.1 [7.3, 9.0]%) versus 59.0 (51.0, 67.0) mmol/mol (7.5 [6.8, 8.3)%) (p=0.024); were less likely to hold a current driving licence, 60% versus 21.2% (p<0.001); and to have previously surrendered their driving licence, 22.2% versus 2.3% (p=0.004). They were also more likely to have had a diabetes-related hospital admission in the previous 12 months, 26.7% versus 7.1% (p=0.004) and to have had at least one day off work/education in the previous 12 months due to hypoglycaemia, 38.1% compared to 2.2% (P<0.001). There was a trend towards those reporting a SH episode being younger at the time they were diagnosed with T1D, 11.0 (8.0, 29.0) years compared to 18 (12, 27) years (p=0.054).

**Table 3:** Characteristics of Subgroups of Severe Hypoglycaemia (SH) in the Past 12 Months.

| Subgroup – number (n) (%) | No Episodes of SH | At least 1 episode of SH | P-Value |
| --- | --- | --- | --- |
| Number (n) – (%) | 157 (84.0) | 30 (16.0) |  |
| Age (years) | 42 (30, 52) | 34 (23.8, 48.5) | 0.161 |
| Female – n (%) | 83 (53.2) | 21 (70.0) | 0.109 |
| Age at Diagnosis (years) | 18 (12, 27.0) | 11.0 (8.0, 29.0) | 0.054 |
| Time Since Diagnosis (years) | 19.6 (11.7, 29.3) | 15.8 (11.1, 28.0) | 0.655 |
| HbA1c (mmol/mol) | 59.0 (51.0, 67.0) | 64.5 (55.7, 75.3) | **0.024** |
| HbA1c (%) | 7.5 (6.8, 8.3) | 8.1 (7.3, 9.0) |  |
| CSII User – n (%) | 73 (46.8) | 7 (23.3) | **0.026** |
| Time on CSII (years) | 5.5 (2.5, 8.2) | 0.7 (0.3, 7.9) | 0.083 |
| isCGM – n (%) | 128 (82.2) | 22 (73.3) | 0.164 |
| If isCGM user, n scans per day | 9.0 (6.25, 12.0) | 9.25 (6.0, 10.0) | 0.789 |
| Time on isCGM (years) | 3.7 (3.1, 4.3) | 4.0 (3.1, 4.5) | 0.428 |
| rtCGM User | 18 (11.5) | 3 (10.0) | 0.168 |
| Hospital Admission in the past 12 months – n (%) | 11 (7.1) | 8 (26.7) | **0.004** |
| Holds driving licence – n (%) | 123 (78.8) | 12 (40.0) | **<0.001** |
| Previously surrendered driving licence – n (%) | 3 (2.3) | 4 (22.2) | **0.004** |
| Unemployed – n (%) | 10 (6.5) | 8 (26.7) | **0.036** |
| Low SIMD Quintile (quintile 1-3) – n (%) | 59 (38.1) | 20 (66.7) | **0.005** |
| Time off work in the past 12 months for hypoglycaemia – n (%) | 3 (2.2) | 8 (38.1) | **<0.001** |
| C-peptide (pmol/L) | 3 (0, 17.8) | 0 (0, 6.9) | **0.04** |
| Average glucose (mmol/L) | 9.9 (8.7, 11.7) | 12.3 (10, 13.6) | **0.021** |
| % Time in Range (3.9-10 mmol/L) | 50 (36, 65) | 38 (25, 55) | 0.053 |
| % Time Below Range (<3.9 mmol/L) | 2 (0, 4) | 2 (0, 5) | 0.873 |
| % Time Above Target (>10 mmol/L) | 46 (32, 63) | 58 (39, 74) | 0.067 |
| % Time Sensor Active | 92 (80, 97) | 78 (67, 87) | **0.009** |

Data presented as median (IQR) unless otherwise specified. Abbreviations: CSII- continuous subcutaneous insulin infusion, isCGM- intermittently scanned continuous glucose monitoring, IQR- interquartile range, rtCGM- real-time continuous glucose monitoring, SIMD- Scottish Index of Multiple Deprivation, SH- severe hypoglycaemia

Participants reporting an episode of SH in the previous 12 months had a significantly lower C-peptide than those not reporting an episode, median (IQR) 0 (0, 6.9) pmol/L versus 3 (0, 17.8) pmol/L (p=0.04) (Figure 3).

**Fig 3.**
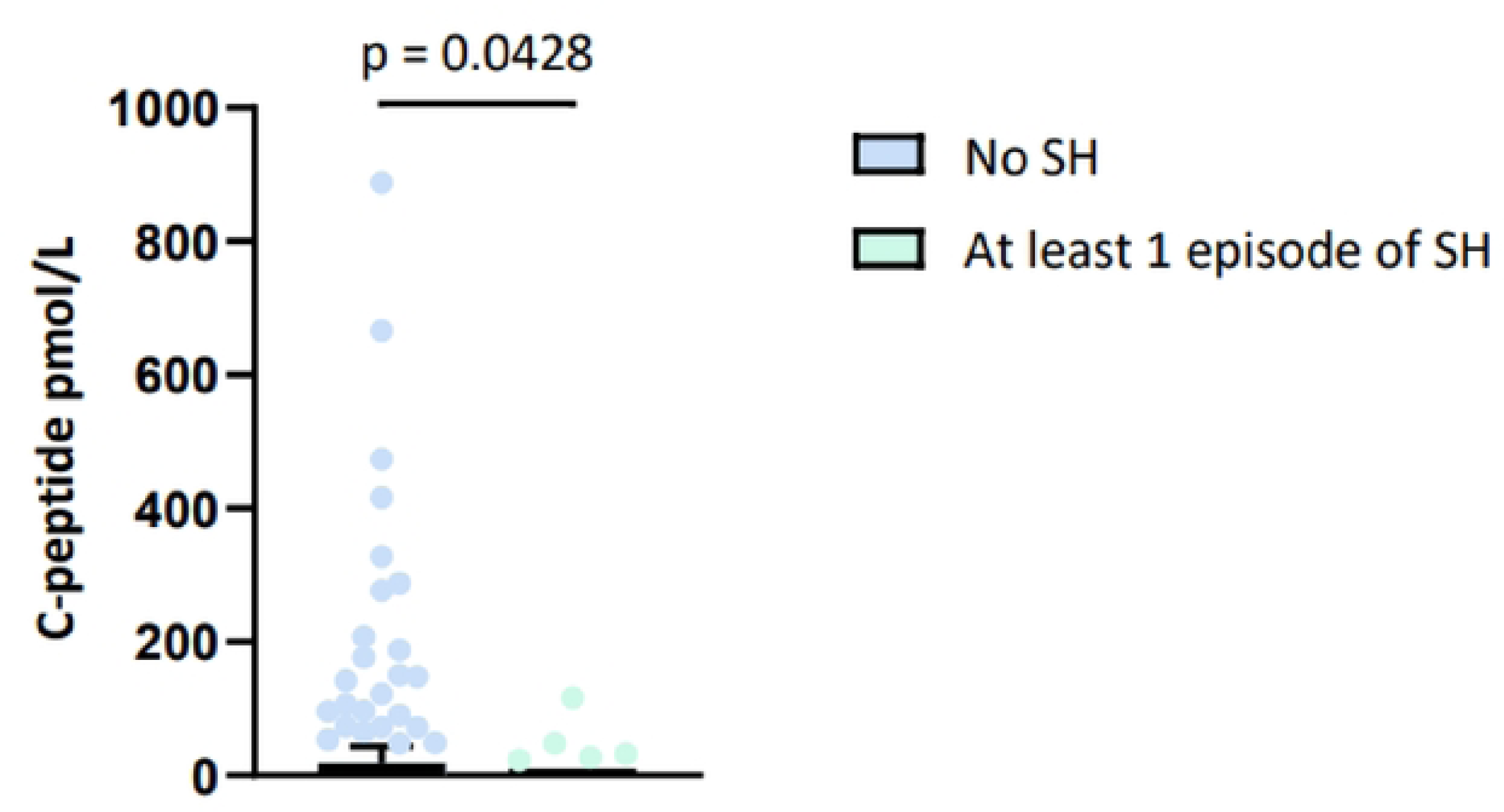
History of Severe Hypoglycaemia (SH) in the past 12 Months and Random C-peptide level.

SH was associated with socioeconomic deprivation. Those reporting an episode of SH were more likely to be unemployed, 26.7% compared to 6.5% (p=0.036), and to come from the most deprived SIMD quintiles, 66.7% compared to 38.1% (p=0.005).

People with a history of SH were more likely to be MDI users, 76.7% compared to 53.2% (p=0.026). There was no group difference in the method for monitoring blood glucose with the majority using isCGM in both the SH group and the group with no history of SH, 73.3% and 82.2% respectively (p=0.164).

Participants with a history of SH had a higher average glucose, 12.3 (10, 13.6) mmol/L compared to 9.9 (8.7, 11.1) mmol/L (p=0.021). There was a non-significant lower TIR in those with a history of SH, 38 (25, 55) % compared to 50 (36, 65)% (p=0.053) and increased TAR, 58% (39, 74%) compared to 46% (32, 63%) (p=0.067). Active sensor time was active significantly lower in those with a history of SH, 78 (67, 87) % compared to 92 (80, 97)% (p=0.009).

## Discussion

In this cross-sectional survey study completed by 189 adults with T1D the prevalence of IAH was 17.4%. This is similar to a Norwegian study by Olsen et al in 2014 which also found a prevalence of IAH of 17% (25). and implies that the prevalence of IAH has not changed in almost 10 years despite advances in technology.

It is, however, a slightly lower rate than reported in a previous study from our centre, which surveyed 518 people with T1D and identified IAH in 19.5% of respondents (6). However, the population characteristics between this study and ours are different. The previous study’s population was younger at the time of completing the survey, median (IQR) age 39 (31–50) years; had a shorter time since diagnosis, 16 (9–24) years and had a higher mean (SD) HbA1c, 68.3 (15.3) mmol/mol (8.4 [1.4]%). Diabetes management amongst respondents in the previous study was also different with no respondents using CSII. This may limit the comparability of these studies. Another recent cross-sectional survey study investigated SH and IAH in CGM users (26). This survey cohort had high levels of technology use, 80% CSII users and 61% HCL users. Despite this they report higher levels of SH and IAH in their cohort, 33% had a Gold score ≥4 and 34.6% had experienced an episode of SH. It may be that a higher proportion of people in their cohort with IAH or a history of SH were on more advanced technologies as a result of these problems. However, it highlights the importance of these high-risk populations being included in future studies of these devices to assess the impact. Similar to our study, this survey found those with a history of SH had higher HbA1c and average glucose levels.

In our study, those reporting IAH were 4 times more likely to report at least 1 episode of SH in the previous 12 months, which is concordant with other studies demonstrating an association between IAH and SH (27). SH is linked to morbidity, mortality and reduced quality of life (QoL), making it an important target for interventions in people with T1D. While there was no difference in the type of insulin or technology used in the IAH subgroups, we did identify that those using MDI were almost 3-times more likely to have had an episode of SH in the previous 12 months. The data supporting a positive impact of diabetes technology on IAH is scant. A recent paper by Ali et al (2023) suggested CGM has reduced the prevalence of IAH, though this premise has been challenged (28). Many studies of insulin technologies do not include people who are at risk of hypoglycaemia, that is, people with a history of SH or IAH, and so it can be difficult to comment on the impact of these devices on the risk of IAH. The lack of representation of these patient cohorts in research studies investigating these devices is a problem. One of the few studies to show improvement in IAH ((10)) found that diabetes education was key, with no difference between technology groups.

The authors acknowledge that the use of the most advanced technologies, such as HCL systems and rtCGM was low in our study, however, this is representative of our local T1D clinic population as reimbursement for isCGM is standard. We did identify a significantly higher proportion of people with normal awareness using a DIY rtCGM system than those with IAH. This is likely due to IAH being a criterion for receiving funded rtCGM in our clinic. Locally, IAH and SH are criteria for referral for more advanced diabetes technologies, however, this work highlights the need to assess patients who are not able to attend clinic using other modalities. It is this sort of data that can influence policy, improve community outreach and aid the development of strategies to help inform and improve access to technology, which may improve uptake and engagement in those difficult to reach lower socioeconomic groups. Such strategies may include reducing barriers to access technologies by offering these to all, increasing provision of information out with the hospital setting and, importantly, peer support. IAH and a history of SH was associated with the most deprived SIMD quintiles. Health disparities exist in T1D (27–29) and previous studies have reported an increased risk of SH in people from more deprived socioeconomic backgrounds (29). However, few studies have linked IAH with socioeconomic status as reported here. Only 5.7% of all respondents in this study were from the most deprived SIMD quintile (quintile 1) and 56.8% were from the least deprived quintiles (quintile 4 and 5). This means that the prevalence of IAH and SH in people from the most socioeconomically deprived areas is likely underestimated in this study. The low proportion of people from the most deprived areas completing this study may in part be due to the questionnaire being administered at a face-to-face clinic: people from the most deprived socioeconomic backgrounds often face more barriers to engaging with health appointments and so the population we have surveyed is not likely to be fully representative of this group. Previous studies, indicate that non-attenders may have poorer glycaemic control (29) and conceivably higher rates of SH and IAH. Although we do not have ethical permission to collect this data we may have underestimated the prevalence of both SH and IAH in the general clinic population. In the IAH group there was a higher proportion of female respondents compared to males, 72.7% versus 52.2%. This may be skewed by the higher proportion of female respondents in the study, 55.7%. this is not a pattern that has been previously reported in studies investigating IAH. As previously noted, a higher proportion of male respondents held a driving licence and a category of licence other than for driving a car alone. Some of these respondents may have had these licence categories as part of their job, which may have made them less forthcoming about their hypoglycaemia awareness status, although confidentiality of the information collected was assured at the time consent was taken.

Contrary to previous studies C-peptide levels in this patient cohort were not associated with CGM glycaemic metrics such as average glucose, TBR and TAR (30, 31). However, in these studies C-peptide levels were higher. We did see a reduced incidence of SH in those with higher detectable C-peptide concentrations as has been reported previously(32) demonstrating the benefit of even very low levels of C-peptide concentrations against hypoglycaemia concordant with studies in islet transplant recipients(33). We did not demonstrate an association between C-peptide and HbA1c in our sample. A previous study investigating the impact of random C-peptide on risk of complications and glycaemic control found a lower HbA1c in participants with a C-peptide >200pmol/L (34).

This study does have limitations: This study may not necessarily be representative of the wider Type 1 diabetes population as people attending the clinic are more likely to be compliant with treatment and prepared to embrace new technologies versus those who do not attend. Also, older people were more likely to complete our survey, which meant that there were few participants with short duration Type 1 diabetes. A further limitation was that the study was completed as a one-off survey and so relied on the recall of participants at a single point in time, however, recall of SH events in the previous 12 months has been shown to be robust (35). This also means that we do not have information about the hypoglycaemia awareness status or history of SH in respondents before they started using diabetes technologies.

The survey was also carried out at a single site and only included participants attending a face-to-face clinic. This may have selected out more motivated and possibly a better controlled cohort than the general clinic population. With this in mind, the authors feel it is important for future studies to reach a wider general clinic population, so that the true extent of these problems can be assessed, and management plans instigated to prevent associated morbidity and mortality.

We also recognise that renal failure is a major risk factor for severe hypoglycaemia. We do not have information regarding renal function in patients. However, intervention with diabetes technology may still positively impact this group and this could be the basis for future work.

## Conclusions

IAH remains a problem for people living with T1D with a prevalence rate of 17.4% in this cohort. In our cohort IAH was associated with a 4-fold increased risk of SH. Both IAH and SH were more prevalent in females and those from a more deprived socioeconomic background and respondents with these problems were more likely to be unemployed. Our study did not identify any difference in the use of diabetes technologies between groups in those who were aware vs those with IAH. However, SH was lower in those using technology.

As has been demonstrated in other studies detectable C-peptide concentrations even at very low levels are protective against SH. Similar to real-world observational studies we found that IAH and SH are associated with higher HbA1c and average glucose levels.

Randomised controlled trials are required to investigate if and how advanced diabetes technologies are beneficial for participants with IAH and, or a history of SH. Since IAH and SH were more prevalent in the most socioeconomic deprived areas, it is important that participants are actively recruited from these groups.

## Data Availability

All relevant data are within the manuscript and its Supporting Information files.

## Acknowledgements

The authors would like to thank the staff and patients of the type 1 diabetes clinic, outpatient department 2, Royal Infirmary of Edinburgh without whom this study would not have been possible.

This study was supported by the Helmsley Charitable Trust (Charity EIN 13-7184401) from whom FB and NB received funding.

